# SIMPLseq: a high-sensitivity *Plasmodium falciparum* genotyping and PCR contamination tracking tool

**DOI:** 10.1101/2025.11.03.25339432

**Authors:** Philipp Schwabl, Jorge-Eduardo Amaya-Romero, Katrina A. Kelley, Paulo Manrique, Sean C. Murphy, Peter D. Crompton, Daniel E. Neafsey

## Abstract

**Background:** Pathogen genotyping via polymerase chain reaction (PCR) amplicon sequencing (AmpSeq) is an informative disease surveillance tool. Several large AmpSeq panels containing >100 multiplexed PCR amplicons have been developed as alternatives to whole-genome sequencing (WGS) methods for the *Plasmodium* spp. parasites that cause malaria, especially for parasite drug resistance tracking and relatedness analysis. However, these large multiplexes often require a costly pre-amplification stage and typically yield sparse data for samples with parasitemia below 10 parasites/μl. Smaller multiplexes optimized for low-parasitemia genotyping have received insufficient methodological work but have the potential to serve multiple important applications. Managing contamination risk during PCR steps represents another key methodological gap that requires attention in the AmpSeq field.

**Methods:** Here we describe a new 6-locus *Plasmodium falciparum* AmpSeq ‘miniplex’ (SIMPLseq) optimized for high-sensitivity analyses that also integrates a contamination detection system based on well-specific inline barcodes applied during first-round PCR (PCR1; in addition to conventional indexing steps in the second-round PCR). We assess panel diversity using publicly available WGS and use mock samples to estimate sensitivity and precision relative to 4CAST, a previously described miniplex. We also create deliberate contamination events to assess sensitivity and estimate unintentional contamination rates during assay application to malaria-infected dried blood spots collected in Mali.

**Results:** SIMPLseq shows high haplotypic diversity *in silico*, distinguishing 96.0% of sample pairs drawn randomly from 12 subnational sample sets. SIMPLseq outperforms 4CAST in sensitivity analyses, achieving 100% average locus detection at ≥0.5 parasites/μl and ≥50% average locus detection at 0.25 and 0.125 parasites/μl, with zero false-positive haplotypes across 25 replicates. Inline barcoding did not significantly affect yield when using a ‘sentinel’ design, whereby one of the six multiplexed PCR1 primer pairs contains the well-specific sequence pair. Sentinel barcoding correctly identified all 24 contaminations introduced deliberately during PCR1 product handling and identified 39 unintentional contaminations in the 1420-sample Malian run.

**Conclusions:** SIMPLseq significantly extends the malaria genomic epidemiology toolkit, employing a simple laboratory protocol based on entirely open-source reagents that is significantly more sensitive and also less costly than most other targeted sequencing protocols. Key use cases for SIMPLseq include recurrent infection classification, polyclonality estimation, and genotypic infection endpoints in intervention efficacy trials.

## Background

Amplicon sequencing (AmpSeq), the deep sequencing of polymerase chain reaction (PCR) products, is a simple, cost-effective, and highly versatile genotyping method with broad utility in infectious disease research and many other fields. In the context of malaria epidemiology, AmpSeq use cases include parasite/vector taxonomic identification (1), transmission intensity evaluation (2), drug/diagnostic resistance surveillance (3), recurrent infection classification (4), and measurement of population connectivity or case importation (5).

In order to maximize information capture, reduce cost, and address multiple malaria use cases at once, several large multiplexed AmpSeq panels have been developed in recent years. Notable examples include SpotMalaria (n = 136 amplicons, via 3 primer pools), which prioritizes drug resistance marker evaluation in the malaria parasite *Plasmodium falciparum* but also contains ‘speciation’ and SNP barcoding-associated targets for diversity analysis (3), AMPLseq (n = 129, via 1 primer pool), which emphasizes both parasite relatedness and drug marker inference (6), and MAD^4^HatTeR (n = 276 (max.), via 2 primer pools), which further incorporates HRP2/3 deletion and vaccine target screens as well as non-*falciparum* infection detection via lactate dehydrogenase gene (7). A key limitation of large multiplex designs is reduced sensitivity, as PCR efficiency often declines due to increasing potential for off-target amplification and primer-primer reaction products when primer diversity is high.

Smaller, ‘miniplex’ AmpSeq designs are therefore of special interest for studies in which low parasitemia infections are common but low false negative rates and high sequencing depth are important to analysis. One important example is presented by malaria intervention efficacy trials in which rigorous malaria strain distinction methods are required to quantify the complexity of infection (COI; number of strains in a single sample) or molecular force of infection (molFOI; number of newly incident parasite strains infecting study participants over time) (4). The active serial sampling designs carried out in these trials, often in highly endemic malaria regions, means that many samples are expected to be parasite-free, and that positive samples will often constitute asymptomatic or pre-clinical infections exhibiting very low parasitemia (<10 parasites per microliter (p/μl) of venous blood) (8). Other malaria use cases that require high genotyping sensitivity while minimizing false-positive signal include entomological surveillance projects, where parasite DNA is recovered at low abundance from mosquito specimens or pools; active cross-sectional surveillance in elimination-phase settings, where remaining cases are biased towards ‘silent reservoirs’; and retrospective studies of archival samples, where sample degradation and depletion often complicate analysis (9). One of the few *P. falciparum* AmpSeq miniplexes currently available is called 4CAST for its combination of CSP, AMA1, SERA2, and TRAP antigen amplicons (6). This 4-plex was applied to profile antigen diversity in samples from an observational longitudinal pediatric cohort study involving >13,000 samples (10). However, the 4CAST design process did not prioritize reaction sensitivity nor did it incorporate a method to detect or minimize DNA contamination (11), an important feature for precision health contexts such as genotypic endpoints for clinical intervention studies.

Contamination is of concern because AmpSeq library construction typically consists of at least two exponential amplification steps, a ‘PCR1’ reaction targeting genomic DNA (gDNA) and a ‘PCR2’ reaction which appends indexing and sequencing adapter components to PCR1 reaction products. Sample plates generally must be physically unsealed and manipulated between these two amplification steps, at which time the reaction’s target regions occur as millions of amplicon copies relative to the original template. Laboratories performing AmpSeq typically attempt to reduce contamination risk through a combination of precautionary measures, including the spatial separation of laboratory spaces used for amplified material, dedicated instruments and reagents per protocol step, and frequent surface sterilization using nucleic acid-degrading agents or ultraviolet light. Such precautions help reduce environmental, experiment-to-experiment carryover, but offer little direct protection against well-to-well contamination (microdroplets or aerosolic transfer) within actively handled multiwell sample plates. The inclusion of negative control wells provides limited further assurance, as these primarily identify major contamination issues affecting multiple wells as opposed to more localized well-to-well transfer events. An additional, commonly applied safeguard is therefore to apply a relatively strict minimum read threshold, e.g., 50 - 350 reads per sample locus (12,13,4), to distinguish true signal from potential contamination artifact. This approach is problematic because it assumes that contamination produces lower read counts than true signal, and because it significantly reduces assay sensitivity, in many cases eliminating the possibility to evaluate low-parasitemia samples or minor strains within polyclonal samples.

A simple potential solution is to incorporate well-specific barcodes directly into the PCR1 primer panel, a method initially developed to help diversify early-cycle base fluorescence and stagger similar cluster signals within Illumina AmpSeq runs (14). Such ‘inline barcoded’ PCR1 primers replace their non-barcoded counterparts at equivalent concentrations, embedding unique identifiers into the resulting amplicons without otherwise altering the overall workflow. In addition to identifying contaminated wells, the approach may also limit the need to set arbitrary minimum read thresholds and enable bioinformatic removal of contaminant reads.

This manuscript evaluates the inline barcoding approach for contamination detection within the context of SIMPLseq, a new and highly sensitive AmpSeq miniplex leveraging ‘<u>s</u>ix informative <u>m</u>alaria <u>p</u>arasite loci’ for *P. falciparum* targeted sequencing. We describe assay strengths and weaknesses encountered during the development process and reflect on lessons learned for future AmpSeq assay selection and design.

## Methods

### Marker selection and diversity analysis

We used the MalariaGEN Pf7k whole-genome sequencing (WGS) dataset (15) to screen for AmpSeq miniplex candidate loci with high diversity across global parasite populations. We measured ‘haplotype_diversity’ (scikit-allel v1.3.13) within subnational sample sets (‘administration level 1’ geographic records in Pf7k) using single-nucleotide (SNP) and insertion-deletion (INDEL) variants which passed Variant Quality Score Recalibration (VQSR) truth sensitivity tranche thresholds and other quality filters previously established by Hamid et al 2023. We excluded variants in regions annotated as ‘Apicoplast’, ‘Centromere’, ‘InternalHypervariable’, ‘Mitochondrion’, ‘SubtelomericHypervariable’, or ‘SubtelomericRepeat’ (15) and used only monoclonal samples (>0.99 F_ws_ as provided by www.malariagen.net/resource/34/) missing less than 30% genome-wide variant calls.

Further evaluation of the ability of candidate loci to distinguish pairwise infection comparisons excluded INDEL variants as well as SNP sites coinciding with INDEL variants or homopolymer tracts ≥5 bp. For each geographic region, we drew 20 random samples with zero missingness across the 8 loci of interest. Random sample draws were not checked for the possible inclusion of highly-related or clonal parasites. We recorded the fraction of all possible pairwise comparisons (i.e., 20 choose 2 per region) for which various single and multi-locus haplotypes could distinguish the samples being compared (i.e., whether haplotype strings mismatched by at least one SNP). We also calculated nucleotide diversity for each 20-sample subset by averaging values of π obtained for 10 kbp windows (‘--window-pi 10000’) using VCFtools v0.1.15.

### Sample material and wet laboratory procedures

To test the laboratory performance of new and previously established AmpSeq assays, we generated mock clinical samples of varying parasitemia from the genotypically distinct, culture-adapted lines *P. falciparum* Dd2 and 3D7. We centrifuged 1 ml blood cell culture containing ring-stage parasites (5% hematocrit, 5% parasitemia) at 500 g for 5 min, retained the dense fraction (100 ul at 50 % hematocrit), and thenre-diluted by a factor of 50x using uninfected whole human blood. The resultant 0.1% parasitemia blood sample (5000 parasites/μl) was then spotted onto Whatman FTA cards and stored at room temperature together with cachets of desiccant beads. For gDNA extraction, a single 6.35 mm diameter circle was punched per bloodspot (handheld puncher) and processed via KingFisher Ready DNA Ultra 2.0 Prefilled Plates (ThermoFisher Scientific) following the buccal swab section of the manufacturer’s protocol. We used the same software script for the KingFisher instrument as provided in LaVerriere et al 2022. Following extraction, we generated 10% 3D7 + 90% Dd2 mock mixtures by combining the corresponding extraction stocks (5000 parasites/μl) at a 1:9 ratio (v/v) and we generated lower mock parasitemias by serial dilution in 2 ng/μl human gDNA (Promega Corporation).

1420 dried blood spot (DBS) samples were obtained from a clinical trial that assessed the efficacy of the anti-malaria monoclonal antibody L9LS in children (following a safety trials in adults) in Kalifabougou and Torodo, Mali (ClinicalTrials.gov ID: NCT05304611) (16). Of the 1420 DBS samples, 712 contained ≥0.1 p/μl, as estimated by a highly sensitive qRT-PCR assay conducted individually (no pooling) at the University of Washington (17). The DBS samples were collected at least every 2 weeks during scheduled visits as well as during unscheduled sick visits during April - October 2022. The trial was approved by the ethics committee at the Faculté de Médecine et d’Odonto-Stomatologie and Faculté de Pharmacie at the University of Sciences, Techniques, and Technologies of Bamako and by the national regulatory authorities of Mali. All participants and/or guardians/parents provided written informed consent. Blood spot storage and gDNA extraction occurred with the same Kingfisher extraction protocol as mock samples above.

PCR1 reactions targeted 1 - 6 loci using 10.5 μl reaction volumes for samples designated only for fragment analysis (Agilent BioAnalyzer 2100 High Sensitivity DNA Kit) and 31.5 μl for samples designated for sequencing analysis. PCR1 reactions used KAPA HiFi HotStart ReadyMix (2x), with reaction inputs added in the same relative quantities in all cases. Primer inputs were added such that each component oligo occurred at 0.7 μM within the final reaction and template input volumes always represented 38% of the final reaction. Specific input values are detailed in the context of 6-locus, ‘SIMPLseq’ amplification at dx.doi.org/10.17504/protocols.io.5qpvodd3zg4o/v1. For all assays except 4CAST, for which we followed a previously established protocol (6), PCR1 thermocycling consisted of an initial incubation step at 95°C (3 min); 29 amplification cycles at 98°C (20 s), 57°C (15 s), and 72°C (30 s); and a final extension step at 72°C (1 min). PCR1 products were then subjected to Exonuclease I digestion (ThermoFisher Scientific) and subsequently diluted 1 to 2 (v/v) in H_2_O. For comparative purposes, we also tried omitting this digestion step (either keeping PCR1 products 100% concentrated or diluting them by 1 to 9 (v/v) in H_2_O). All PCR2 reactions used 5 μl KAPA HiFi HotStart ReadyMix, 3 μl PCR1 template (digested or otherwise), 2 μl H_2_O, and 2.2 μl unique dual index (10 μM input for a final concentration of 1.8 μM). PCR2 cycling consisted of an initial incubation step at 95°C (3 min); 10 amplification cycles at 98°C (20 s), 65°C (30 s), and 72°C (30 s); and a final extension step at 72°C (1 min).

PCR2 products were subsequently pooled and subjected to left-sided (small fragment) clean-up using AMPure XP beads. When simultaneously processing PCR2 products representing distinct multiplexes, pooling and bead clean-up occurred separately for each multiplex. We diluted cleaned pools (generally 1 to 99 (v/v) in 10 mM Tris-Cl) for Bioanalyzer visualization and performed final quantification using the KAPA Library Quantification Kit (Roche). For sequencing of mock sample sets in-house, pools were brought to 4 nM using 10 mM Tris-Cl, denaturated with an equal volume of 0.2 N NaOH, and diluted to 14 pM using hybridization buffer (Illumina HT1). Mock sample sequencing was performed using 500-cycle Illumina MiSeq (Reagent Kit v2) with the addition of 20% PhiX. In order to standardize yield comparisons between multiplexes, multiplexes were sequenced with equivalent initial PCR input template configurations, either on the same run or using a common calibrator library (AMPLseq) included at 10% concentration on separate runs. For clinical sample sequencing (L9LS study), libraries were sent to the Broad Institute Walk-up Service for paired-end NovaSeq SP, likewise using 500 paired-end cycles and 20% PhiX.

### Inline barcoding at PCR1

To detect cross-well contamination events during library preparation involving PCR, we tested the use of SIMPLseq PCR1 primers containing inline barcodes between the target-binding region (3’) and the Nextera adapter sequence (5’). We provide an illustration of this concept in the Results section. For initial read yield testing of SIMPLseq multiplexes containing inline barcodes at one or multiple loci, we equipped PCR1 forward primers with inline barcode sequence CTTGGCAT and PCR1 reverse primers with inline barcode sequence CGTGGTAA. This barcode pair was previously identified as a good representative of reaction yield among 96 distinct barcode combinations applied to all SIMPLseq primers in a combinatorial format (a distinct forward barcode in each column combined with a unique reverse barcode in each row) as detailed in Supplementary Table 1. In further assay development pursuing a single-locus tagging format (applying barcodes only to the KELT primer pair, instead of to all 6 primer pairs within the SIMPLseq multiplex), we additionally tested shorter (3 bp) barcode pairs (GTA + CAT, ATG + CAT, and ATG + TCA) relative to a subset of longer (8-11 bp) barcode pairs from Supplementary Table 1 (A04, D03, F08, F12, G05, and G10). Wider application of the single-locus tagging approach to clinical samples from Mali applied all barcode pairs from Supplementary Table 1 (i.e., 96 different KELT-tagged primer pairs, manually combined from individual tubes).

### Inline barcode detection within FASTQ files

The detection of inline barcodes was conducted directly on FASTQ files, prior to the microhaplotype denoising workflow described in the next section. The detection workflow is available in GitHub (18) and can be run with a graphical user interface via the cloud-based Terra data analysis platform (19). The pipeline first applies a minimal amount of 3’ quality-trimming (base calls with Phred scores below 5) and then trims any present adapter sequences using Trim Galore v0.6.6. Reads shorter than 20 bp after trimming are discarded and the remaining forward and reverse reads, including inline barcoded reads, are merged using the command bbmerge.sh from BBMap v39.01 (default parameters). The resultant merged FASTQ files are used to quantify matches between inline barcodes within reads to a text file of barcode pairs expected per sample. In the case of long barcodes (≥8 bp), we allowed up to two mismatches between query and reference strings to consider a match. For shorter barcodes, we did not allow any mismatches to occur. In the context of mock contamination experiments (quantifying the occurrence of 3 bp barcodes in amplicons containing Dd2 vs. 3D7 insert sequences), we searched the merged FASTQ files (grep) for perfect matches to either reference FASTA (including also primer binding sites) and tabulated how often matches were bounded by expected/unexpected barcode pairs.

### Sequence read denoising

To denoise PCR and sequencing errors from sequencing reads, we first stripped FASTQ files of inline barcode occurrence using cutadapt v3.4 (‘-g barcodes.fasta’, where barcodes.fasta represents an anchored list (e.g., ^GTA etc.) of all barcodes used in the sample set). We then applied the publicly available malaria amplicon processing pipeline (20) using previously described default settings (6). Briefly, the pipeline first discards read-pairs missing primer sequences (max mismatch = 2) and then trims primer sequences from remaining reads (TrimGalore v0.4.5). It subsequently performs quality processing within DADA2 v1.14.0, setting maximum expected errors (maxEE) to c(5,5), hard 3’-clipping (trimRight) to c(2,2), and soft 3’-clipping (truncQ) to c(5,5). Next, read denoising occurs using standard pooled processing (pool = T) with de novo error model construction from the input data (SELF_CONSIST = T with MAX_CONSIST = 10). The evidence threshold for new read variant assignment (OMEGA_A) is set to 10^-120^. Denoised sequences are then merged (no mismatches allowed) to microhaplotypes and aligned to 3D7 reference sequences using the ‘pairwiseAlignment’ function in Biostrings v2.60.0. Using very permissive edit distance (number of SNP differences with respect to 3D7) thresholds per locus (median 49.3), excessively divergent microhaplotypes are discarded. Microhaplotypes exceeding 10% INDEL distance to 3D7 are also discarded. Finally, SNPs within homopolymers of length >5 bp are masked and microhaplotypes are abbreviated into a compact ‘pseudo-CIGAR’ format. Unless otherwise stated, we further filtered pipeline output by discarding microhaplotypes belonging to sample loci supported by <100 total read-pairs (‘minimum locus depth filter’) as well as microhaplotypes supported by <1% total read-pairs within a sample locus. Microhaplotypes were also required to occur in at least 2 samples if they did not represent the major allele in any sample (i.e., no ‘singleton’ minor alleles).

## Results

### Candidate panel exploration in silico

To initiate our miniplex assay development, we drew from a previous compilation (6) of high-diversity *P. falciparum* monoplex amplicons from multiple studies (3,21–24). We ranked 128 previously established ‘AMPLseq’ targets (119 of which had been specifically tailored towards multiplexed reaction using the GTseek commercial service (25)) based on regional haplotype diversity analysis and focused on the 80^th^ percentile for miniplex target selection (Figure 1A). We pre-screened target candidates for amplification efficiency by assessing PCR2 product size distributions generated by previously designed primer pairs. Fragment traces indicated high off-target amplification for AMA1 (PF3D7_1133400) and SERA2 (PF3D7_0207900) (members of the previously published 4CAST assay, which is composed of primers designed for monoplex amplification), both in monoplex amplification as well as when multiplexed with the other 4CAST loci CSP (PF3D7_0304600) and TRAP (PF3D7_1335900) (Supplementary Figure 1). High off-target concentrations (especially near 190 bp post-PCR2) persisted across various modifications of the original 4CAST protocol and could not be sufficiently ameliorated via bead-based clean-ups without compromising target peaks (Supplementary Figure 2). Especially in the case of low parasitemia samples, fragment traces indicated higher amplification specificity for 4CAST without AMA1 or SERA2, i.e., for CSP + TRAP, also with the inclusion of up to 4 additional loci (WDCP (PF3D7_1410300), KELT (PF3D7_1475900), SERA8, (PF3D7_0207300), and SURFIN 4.2 (PF3D7_0424400)) in the six-plex reaction we call SIMPLseq (Supplementary Figure 3).

**Figure 1.**
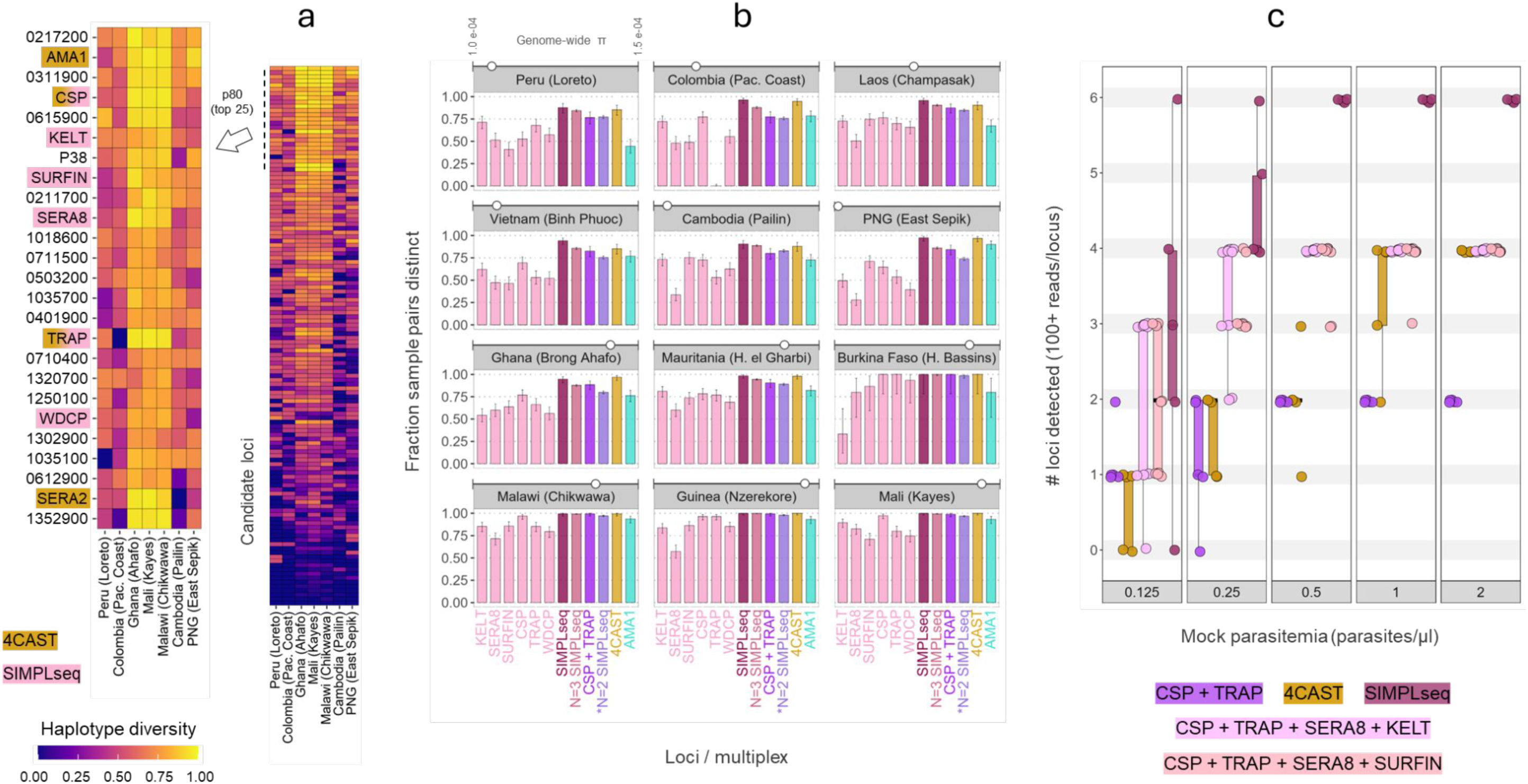
Panel selection based on haplotype diversity and locus recovery at low parasitemias. **a)** In the right heatmap, loci from the previously established AMPLseq panel are sorted by mean haplotype diversity within subnational sample sets (20 random samples taken from each of 7 different ‘administration level 1’ geographic records in Pf7k). The top 25 loci are expanded at left, labeled either with gene ID (minus PF3D7 prefix) or common shorthand. Gold and pink highlighting indicates loci included in 4CAST and SIMPLseq, respectively. **b)** The ability of SIMPLseq markers to distinguish random subnational sample pairings, either based on individual loci or based on multi-marker combination, is shown relative to 4CAST and AMA1. On the x-axis, ‘SIMPLseq’ markers refers to the full 6-locus panel, and ‘N=2 SIMPLseq’ denotes all 2-locus combinations of SIMPLseq markers excluding CSP and TRAP, which are plotted separately. Genome-wide nucleotide diversity is additionally shown via white circle at the top of each panel. Panels represent analysis of random N=20 sample set except in the case of Burkina Faso, for which only 6 samples were available. **c)** Parasitemia levels (x-axis bins) represent cultured ring-stage Dd2 parasites mixed with whole human blood prior to DNA extraction. Boxplots represent median and interquartile ranges for the number of loci detected with at least 100 reads using different amplicon panels.

We assessed the ability of the SIMPLseq marker set to distinguish random monoclonal sample pairings *in silico* with respect to 4CAST and AMA1, the most diverse locus in 4CAST (Figure 1B). SIMPLseq and 3-locus SIMPLeq subsets (20 possible 3-locus combinations) generally performed similarly to 4CAST and, especially in lower transmission settings (e.g., regional foci in South America and Asia), outperformed AMA1. Across all tested regional contexts, the average percentage of sample pairs resolved as distinct was 96.0% (95% confidence interval (CI): 95.0 - 96.8%) for SIMPLseq; 91.4% (CI: 91.1 - 91.7%) for 3-locus SIMPLseq; 94.2% (CI: 93.1 - 95.2%) for 4CAST; and 94.2% (CI: 77.4 - 81.0%) for AMA1. At the individual locus level, however, the SIMPLseq markers generally showed reduced ability to distinguish sample pairs relative to AMA1. The highest average pairwise distinction rate among SIMPLseq loci was observed for CSP (78.5%, CI: 76.6 - 80.2%) and the lowest was observed for SERA8 (53.9%, CI: 51.7 - 56.0%).

### SIMPLseq and alternate panel sensitivity and precision

We proceeded to evaluate SIMPLseq sensitivity relative to 4CAST by sequencing libraries prepared using 12 μl *P. falciparum* (strain Dd2) DNA extracts representing parasitemia levels between 0.125 and 2 parasites/μl in whole human blood. Comparisons also included SIMPLseq subset reactions (CSP + TRAP + SERA8 + KELT; CSP + TRAP + SERA8 + SURFIN; and CSP + TRAP) given possible tradeoffs between multiplex size and sensitivity limits. Full SIMPLseq achieved 100% (6 of 6) locus detection at ≥ 0.5 parasites/μl for all 5 replicates (Figure 1C) but the mean number of loci detected dropped to 4.4 at 0.25 parasites/μl and 3 at 0.125 parasites/μl. 4CAST showed lower sensitivity, failing to achieve complete locus recovery at 0.5 parasites/μl, with a mean of 2 loci detected among its 5 replicates. The mean number of 4CAST loci detected dropped to 1.6 at 0.25 parasites/μl and 0.6 at 0.125 parasites/μl, without detection of AMA1 or TRAP at these two lowest parasitemia levels. The CSP + TRAP duplex showed similar absolute locus counts relative to 4CAST at 0.5, 0.25, and 0.5 parasites/μl. Absolute locus counts for the 4-locus SIMPLseq subsets generally fell in between those of the CSP + TRAP duplex and the full SIMPLseq panel at all parasitemia levels assessed.

Next we verified genotyping precision (% detected haplotypes showing correct Dd2 reference sequence per locus) for each sequenced panel. Under default filtering conditions (excluding minor alleles supported by <1% read prevalence within a sample locus and no singleton minor alleles), all 25 replicates per panel showed exclusively the expected Dd2 haplotypes. Removing the singleton filter (Supplementary Figure 4), SIMPLseq precision remained 100% for WDCP, SURFIN, and SERA8, but decreased to an average of 98% for KELT and TRAP (for each locus, 1 of 25 replicates showed a minor haplotype in addition to the correct call) and 94% for CSP (3 of 25 replicates showed a minor haplotype in addition to the correct call). Similar precision variation was observed with the 2- and 4-locus SIMPLseq subsets (94-100% per locus), but not with 4CAST, for which precision remained 100% in the absence of singleton filtration. These variations in precision correlated negatively to read depth (p = 1.9 e-04) but did not vary by locus in a mixed effect model (precision ∼ totreads + marker + (1 | assay)) with Bonferroni correction.

### Adding inline barcodes to SIMPLseq PCR1

To enable contamination detection during the PCR stages of AmpSeq library preparation, we added ‘inline barcodes’ to the primers used for SIMPLseq PCR1 amplification of the KELT locus. These short (3-12 bp) sequences are placed between the PCR1 primer target-binding sequence (3’) and the adapter sequence (5’) required for primer binding during PCR2 (Fig 2A top). For cost efficiency, inline-barcoded primers can be implemented in a combinatorial design, where each row on a sample plate shares the same forward primer and each column shares the same reverse primer, but each well represents a distinct barcode pair (and dual-uniqueness occurs for wells that do not share the same column or row). The PCR2 reaction remains a standard indexing reaction (Fig 2A bottom), introducing a second pair of sample identifier sequences (generally unique dual indices as opposed to a combinatorial design) and the adapter components (P5 and P7) required for amplicon attachment to the Illumina flow cell.

**Figure 2.**
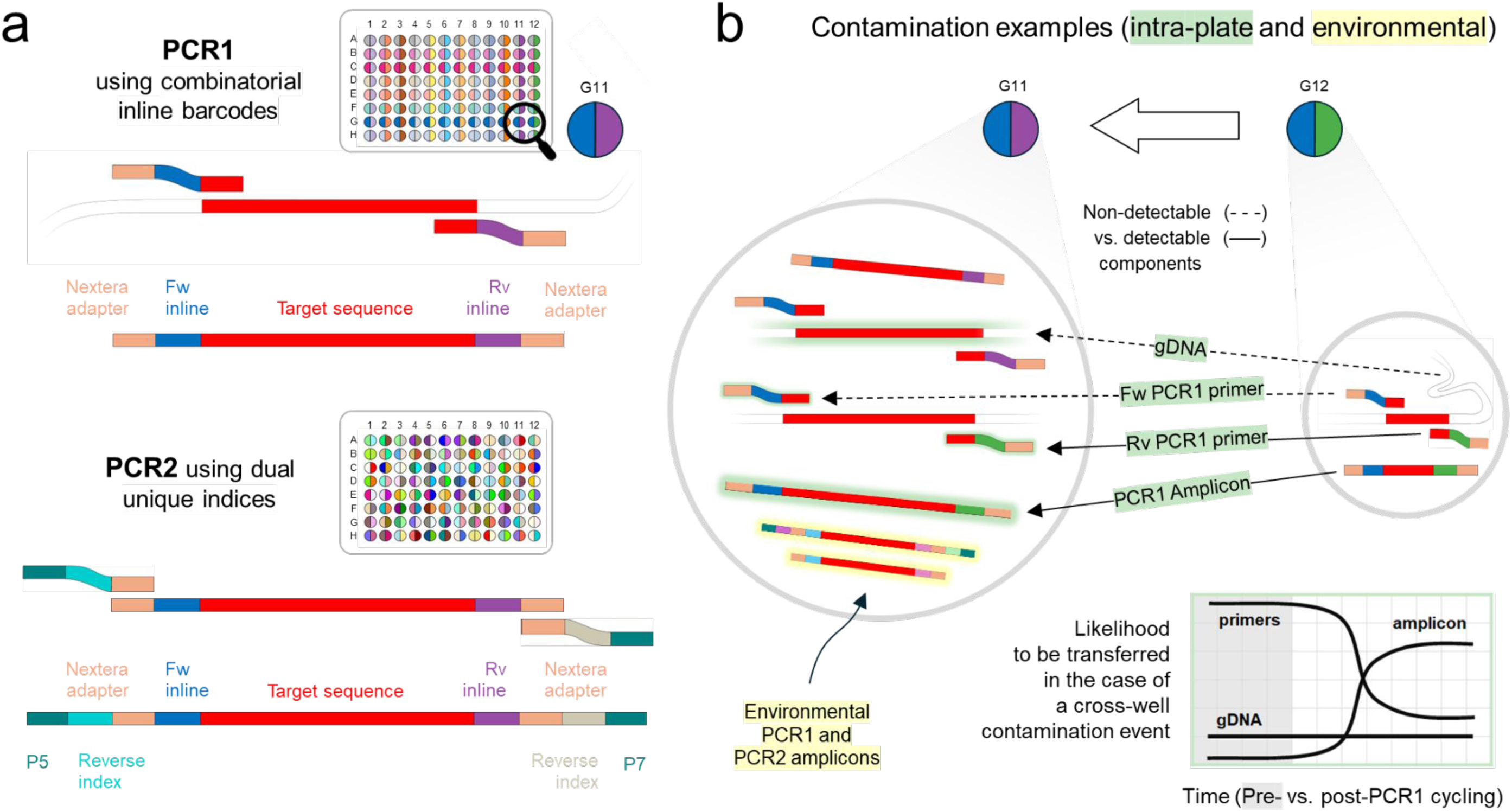
Schematic of PCR amplification steps when inline barcodes are incorporated into the first-round reaction. **a)** AmpSeq workflows very commonly employ a PCR1 reaction in which primer pairs contain universal 5’ tails (see tan segments) in order to create binding sites for subsequent PCR2 reaction. These tails generally also function as sequencing primer binding sites. Inline barcodes can be additionally incorporated within the PCR1 primers. For practical reasons, a combinatorial design can be used where barcodes on forward primers are identical within plate rows and barcodes on reverse primers are identical within plate columns, yet wells remain unique when considering the full barcode pair. The generation of PCR1 amplicons from genomic DNA (gDNA) is illustrated based on the use of primers from well G11 (represented by blue and purple) in a combinatorially arranged primer plate. The PCR2 reaction uses PCR1 products as input, adding sample-specific indices and flow cell adapter sequences (P5/P7) to each 5’ end. PCR2 is the same regardless of whether inline barcoding has been implemented for PCR1. The indices used in PCR2 are generally fully unique (no sharing within rows or columns) in order to preclude index hopping artefacts. **b)** Hypothetical contamination affecting recipient well G11 (large circle) at the PCR1 stage (during plate set-up or upon opening after cycling) are shown. For example, G11 may receive cross-well contamination (see molecules with green glow) from adjacent donor well G12 (small circle). The contaminating molecules may comprise gDNA, PCR1 forward primer, PCR1 reverse primer, or PCR1 product. Using a combinatorial barcoding format, contamination involving G12 forward primer or G12 PCR1 product should be detectable via mismatch to the expected barcode pair. The contamination would be undetectable if it were to exclusively involve gDNA (i.e., no concomitant transfer of mismatching barcodes), but this is not considered likely based on expected molarities. As illustrated in the schematic at bottom right, PCR1 products are expected to dominate wells once cycling is underway, meaning that cross well contamination events involving only undetectable gDNA and not concomitant amplicon (or forward primer) are unlikely to occur. Another contamination scenario involves ‘environmental’ sources (see yellow highlighting), i.e., PCR1 or PCR2 products from other sample plates, laboratory instruments, or reagents used in inline-barcoded AmpSeq workflows may enter the current well. Such events are expected to be detectable in the majority of cases.

The inclusion of inline barcodes during PCR1 means that cross-well contamination events during and post-PCR1 reaction set up are likely to involve the detectable transfer of extrinsic, ‘contaminant’ barcodes to affected wells, either in the form of unincorporated primers or of amplified DNA. Effective contamination is only expected to occur without a detectable barcode signal if the transferred material involves exclusively genomic DNA, i.e., without the simultaneous transfer of reverse primers or amplicons in the case of the G12 to G11 contamination example illustrated in Figure 2B. Such silent contamination is expected to become exceedingly unlikely over the course of an AmpSeq protocol.

We decided on adding the inline barcoding feature to KELT primers as opposed to all SIMPLseq primers based on an initial sequencing test showing that complete tagging reduces read yield and locus detection rate as parasitemia declines (Figure 3). Comparing completely tagged vs. untagged SIMPLseq, we observed a 66% reduction in mean reads per detected locus at 4 p/μl (Tukey HSD p = 8.71 e -5) and a 27% reduction trend in mean number of loci detected at 0.5 p/μl. Meanwhile, single-locus tagging of KELT did not significantly affect read-depth per detected locus at 4 p/μl or show a reduction trend for mean number of loci detected at 0.5 p/μl. Tagging of either CSP or SURFIN also met these criteria, but average read depth trended 21% lower than for the KELT-tagged multiplex (t-test p = 0.016 without multiple testing correction).

**Figure 3.**
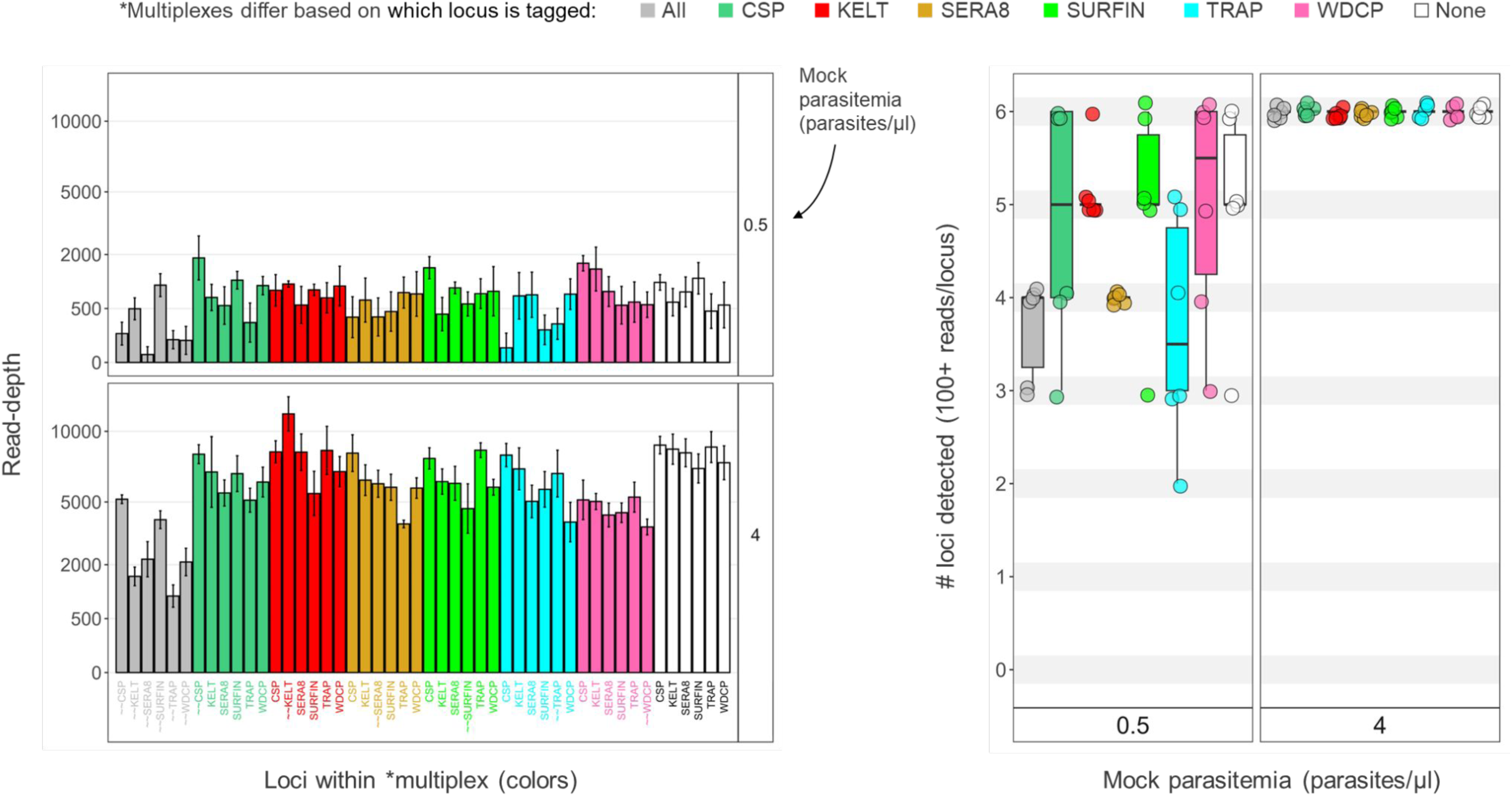
SIMPLseq sensitivity when applying inline barcodes to all six loci or to a single sentinel locus. On the left, the read-depth for each locus within seven different SIMPLseq reactions (fill colors) is shown on the y-axis. Reactions differ based on whether all (grey), none (white), or one of the six SIMPLseq loci (see color key) were targeted using inline barcoded primer pairs. X-axis labels for loci targeted using barcoded primer pairs are prefixed with ‘∼∼’. PCR1 template consisted of Dd2 DNA, either at 0.5 p/μl (upper panel) or 4 p/μl (lower panel). The right plot further compares the different SIMPLseq reactions based on number of loci detected with at least 100 reads, with the two parasitemia panels arranged left to right.

We also further verified KELT-tagged SIMPLseq sensitivity by assessing minor strain detection rate in mock strain mixtures consisting of 10% 3D7 and 90% Dd2 at total parasitemia levels between 0.5 and 64 parasites/μl in whole human blood. We lowered the minimum locus depth filter to 10 for this analysis. No statistical differences occurred between tagged (n = 12 per parasitemia level) and untagged replicates (n = 6 per parasitemia level) (Supplementary Figure 5A). At 64 p/μl, 3D7 alleles were detected for all 6 SIMPLseq loci in all replicates of the KELT-tagged multiplex and all replicates of the untagged multiplex. Mean 3D7 allele detection dropped to 5.0 [1.6 sd] (tagged) and 5.8 [0.4] (untagged) at 16 p/μl, 4.3 [0.8 sd] (tagged) and 2.83 [1.6] (untagged) at 8 p/μl; 1.6 [1.1] (tagged) and 1.3 [1.0] (untagged) at 2 p/μl; and 0.2 [0.4 sd] (tagged) and 0.5 [0.5 sd] (untagged) at 0.5 p/μl. Half of the tagged replicates in these sensitivity tests involved a shorter barcode length than previously tested (3 bp instead of 8-11 bp) barcode types, but no statistical differences in 3D7 allele counts between short vs. long barcode types was observed. Relative read counts for detected 3D7 alleles also appeared consistent with the expected value (10%) for parasitemia levels ≥ 2 p/ul but dropped to 1.9% (tagged assay median) for 0.5 p/μl (Supplementary Figure 5B).

### Identification of deliberately contaminated wells by inline-barcoding KELT

We tested the ability of KELT-barcoding to identify contaminated wells using a PCR1 reaction plate design in which templates (Dd2 gDNA vs. 3D7 gDNA or water) occurred in alternating rows (Supplementary Figure 6). We then deliberately introduced contamination at a different SIMPLseq protocol step, each event affecting a different sample row (i.e., no successive contamination events affecting the same row). These deliberate contaminations involved the pipetted transfer of 0.3 ul from ‘donor’ wells for which initial PCR1 input template was Dd2 gDNA into ‘recipient’ wells for which initial PCR1 input template was either 3D7 (wells 1 – 6) or water (wells 7 – 12). Comparing intended donor and recipient wells, barcoding was either singly unique (i.e., one of the two barcodes used for each sample also being used for the other sample) or dually unique (both barcodes distinct) between intended donor and recipient wells (Supplementary Figure 6). This set-up was meant to enable contamination detection via Dd2-matching (in denoised microhaplotypes, no minimum locus depth filter) as well as via contaminant barcode occurrence (in FASTQ files) – see Methods.

Contamination event 1 (row B to C) was performed just prior to sealing for PCR1 amplification (just prior to step 4 in <u>Supplementary Text 1: SIMPLseq protocol</u>), i.e., within the plate containing fully prepared PCR1 reaction mixes. At this stage, each well should contain primers and unamplified template as illustrated in Figure 4A (left). Event 2 (row D to E) was performed just after unsealing PCR1-amplified products (just after step 4 in <u>Supplementary Text 1: SIMPLseq protocol</u>). At this stage, each well should contain primers, unamplified template, as well as target amplicons (Figure 4B, left). Event 3 (row F to G) was performed just prior to sealing for PCR2 amplification; however, instead of within-plate transfers as in Events 1 and 2, the donor samples were taken from the diluted PCR1 digestion product plate and added into the fully prepared PCR2 reaction mixes (i.e., repetition of step 11 in <u>Supplementary Text 1: SIMPLseq protocol</u>). In this scenario, donor wells should contain unamplified template and target amplicons but PCR1 primers should have been eliminated by the prior ExoSap digestion step. Recipient wells should contain PCR2 primers, unamplified template, and target amplicons (Figure 4C, left).

**Figure 4.**
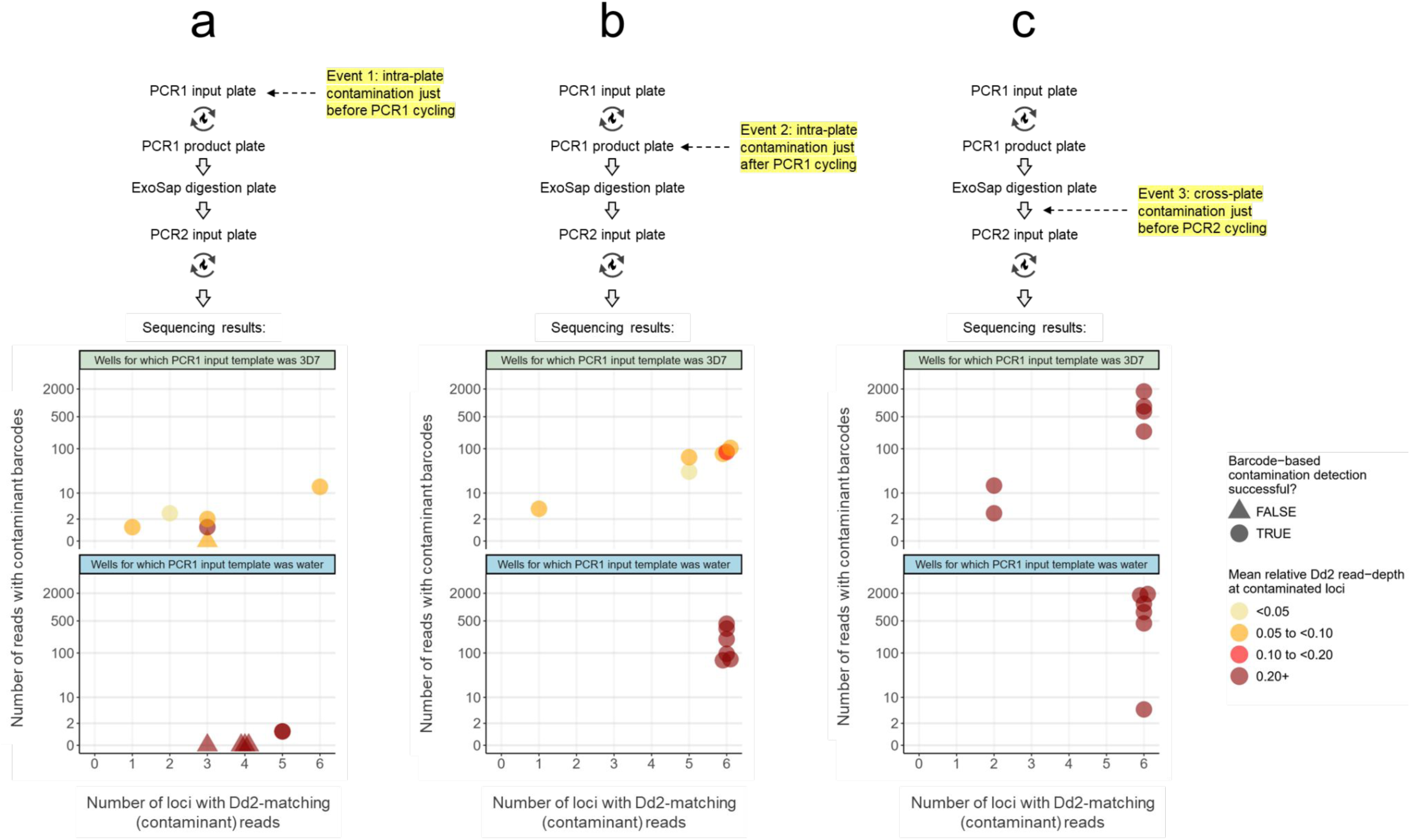
Deliberate contamination experiment: ability to detect contaminated wells using an inline-barcoded sentinel locus. Figures a-c indicate sequencing results corresponding to three deliberate contamination events (12 recipient wells per event) enacted during the SIMPLseq workflow (just before PCR1 cycling, just after PCR1 cycling, and just prior to PCR2 cycling – see yellow highlighting in the upper protocol plots). Contaminations involved 0.3 μl sample transfer from donor wells for which Dd2 DNA was used as PCR1 input template. Recipient wells had either 3D7 DNA (see top panels with green title bar) or water (bottom panels with blue title bar) used as PCR1 input template. Each point represents a recipient well. X-axes indicate effective contamination, i.e., the number of loci for which Dd2-matching reads were detected (at min. xx depth) in the recipient well. Y-axes (square-root scale) indicate contamination detectability via the inline-barcoding approach, i.e., the number of KELT read-pairs with contaminant barcodes detected in the recipient well. Points are colored based on mean relative Dd2 read-depth at contaminated loci. Points are given a triangle shape when inline barcoding-based contamination detection failed, i.e., Dd2-matching reads were detected in the recipient well but contaminant barcodes were not.

Event 1 (intra-plate contamination just prior to PCR1 cycling) led to the occurrence of Dd2-matching reads at an average of 3.0 loci for recipient wells representing 3D7 as initial input (wells 1-6) and 4.2 loci for wells representing water as initial input (7–12) (Figure 4C, right top). Dd2 read signal constituted an average of 8.4% (wells 1-6) and 100% (wells 7-12) of the total strain signal (denoised read-pairs for 3D7- and Dd2-matching microhaplotypes) per affected locus. Identification of Dd2-contaminated wells via contaminant barcode sequence detection occurred successfully for 7 of 12 wells.

Event 2 (intra-plate contamination just after PCR1 cycling) led to slightly higher Dd2-matching read signal, occurring at an average of 4.8 loci for wells 1-6 and 6.0 loci for wells 7-12, and averaging 6.3% and 100% (respectively) of the total strain signal per affected locus (Figure 4C, right middle). Unlike for event 1, contaminated well identification via contaminant barcode detection occurred robustly for all wells, though detection counts trended higher for wells 7-12 (203 barcodes on average) than for wells 1-6 (61 barcodes on average; T-test p = 0.076).

Event 3 (inter-plate contamination just before PCR2 cycling) showed yet higher Dd2-matching read signal than event 2 (Figure 4C, right bottom). Dd2-matching reads occurred at an average of 4.7 loci for wells 1-6 and 6.0 loci for wells 7-12, averaging 29.7% and 100% (respectively) of the total strain signal per affected locus. Contaminant barcode counts also averaged higher for event 3 (805) than for event 2 (132; T-test p = 0.008). Event 3 showed similar contaminant barcode counts between wells 7-12 (1022 barcodes on average) and 1-6 (587 barcodes on average; T-test p = 0.318).

### Potential to clean up contaminant signal for KELT?

We next evaluated whether wells identified as contaminated via contaminant barcodes can be filtered of contaminant signal rather than discarded entirely from further analysis. While such filtering is not directly applicable to a multi-locus panel in which inline-barcoding has only been implemented for a single locus, it would be valuable for single-locus assays, e.g., targeting AMA1.

Within FASTQ files, we therefore examined the relative abundance of Dd2-matching contaminant reads that contain contaminant barcodes vs. those that do not (Figure 5). The latter ‘silent’ form of contamination can occur if contaminant gDNA is amplified by PCR1 primers belonging to the recipient well (creating amplicons with ‘resident’ barcodes yet foreign insert DNA). For Event 1 (Dd2-associated contamination just prior to PCR1 cycling), silent Dd2 reads were preponderant, representing an average of 99.30% of all Dd2 reads and 7.13% of total reads for recipient wells 1-6 (for which 3D7 gDNA was used as initial PCR1 template). For water-associated wells 7-12, Event 1 silent Dd2 signal represented an average of 99.70% of total reads (all of which were Dd2). In the case of contamination after PCR1 cycling (Event 2 and 3), however, silent signal appeared extremely rare. For Event 2, silent Dd2 signal represented an average of 0.28% of all Dd2 reads and 0.04% of total reads for recipient wells 1-6. Event 2 silent Dd2 signal for wells 7-12 represented an average of 1.61% of total reads (all of which were Dd2). For Event 3, silent Dd2 signal represented an average of 1.74% of all Dd2 reads and 0.45% of total reads for recipient wells 1-6. Event 3 silent Dd2 signal for wells 7-12 represented an average of 0.42% of total reads (all of which were Dd2). Absolute read counts for silent contamination at Events 2 and 3 were likewise very low (0 to 18 reads per well), especially when donor-recipient well pairs represented dually-unique inline barcode pairs (1 silent read-pair in G12; all other wells zero). Considering the typical evidence thresholds that researchers apply in microhaplotype analysis (e.g., requiring minor haplotypes to be supported by at least 1% of reads within a locus), the above results suggest that sequencing yield for an inline-barcoded locus in a parasite-positive sample is unlikely to contain any undetectable contaminant reads originating from post-PCR1 cycling contamination events. This means that users have the opportunity to confidently salvage all non-contaminant reads at the barcoded locus if they do not wish to repeat PCR and sequencing for a contaminated sample.

**Figure 5.**
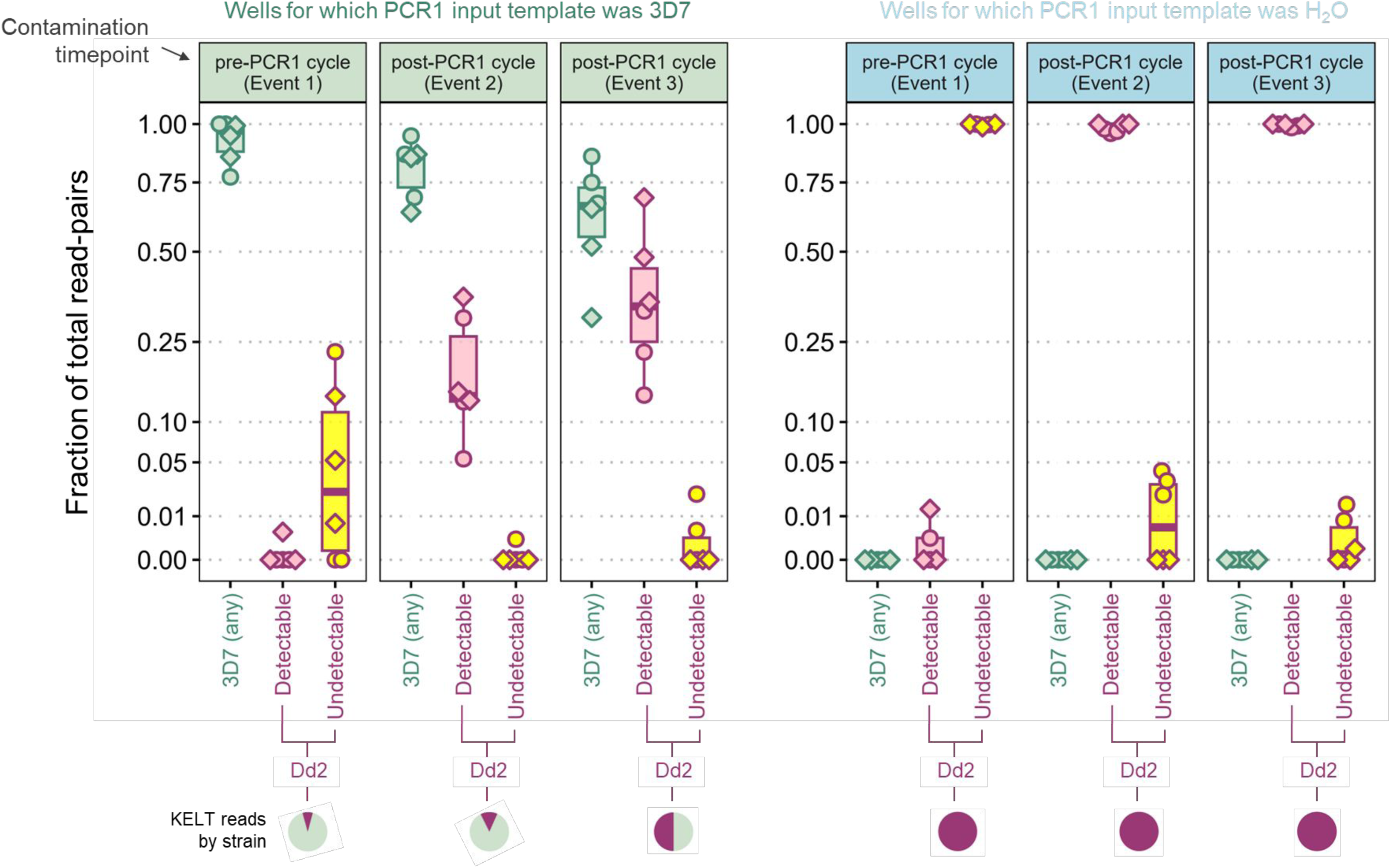
Deliberate contamination experiment: ability to detect contaminant read-pairs at the inline-barcoded sentinel locus. Each of the two plots is composed of three panels corresponding to the three deliberate contamination events enacted during the SIMPLseq workflow (just before PCR1 cycling, just after PCR1 cycling, and just prior to PCR2 cycling – see also yellow highlighting in Fig. 4). Contaminations involved 0.3 μl sample transfer from donor wells for which Dd2 DNA was used as PCR1 input template. Recipient wells had either 3D7 DNA (see panels with green title bar) or water (panels with blue title bar) used as PCR1 input template. Each event involved 12 recipient wells (points). Using boxplots representing median and interquartile ranges, each panel indicates the fraction of total KELT read-pairs (square-root scaled y-axis) belonging to one of three categories: Category 1 (green boxplots at left): insert sequence matches 3D7 (i.e., the sequenced amplicon is of non-contaminant template origin). Category 2 (pink boxplots in middle): insert sequence matches Dd2 (i.e., the sequenced amplicon is of contaminant template origin) and contaminant barcodes occur on the read-pair. This contamination was ‘detectable’ (see x-axis) via the inline-barcoding approach. Category 3 (pink boxplots at right): insert sequence matches Dd2 (i.e., the sequenced amplicon is of contaminant template origin) but contaminant barcodes do not occur on the read-pair. This contamination was ‘undetectable’ via the inline-barcoding approach. Points are given a diamond shape when representing a recipient well for which the designated inline barcode pair is dually unique with respect to the donor well’s designated inline barcode pair (see plate configuration in Supplementary Fig. 6). Pie charts at bottom further help visualize insert identity by strain.

### Unintentional contamination rates

Finally, we assessed the rate at which unintentional contamination occurred during the SIMPLseq protocol (carried out by an expert in sterile technique). In the deliberate contamination experiment described in the previous section, 2 of 42 (4.8%) wells that were not intended to receive contamination (asterisked in Supplementary Figure 6) did in fact unintentionally receive 1 contaminant read-pair each. These 2 wells were intended to amplify Dd2 template, and the contaminant read-pair found within each well contained Dd2-matching insert sequence, suggesting that the unintentional contamination involved only contaminant primer (as opposed to contaminant gDNA or amplicon molecules representing 3D7). In the second reaction plate involving inline barcoded primer pairs (the plate which was used to assess minor strain detectability within strain mixes further above (Supplementary Figure 5A)), 5 of 96 wells also experienced unintentional contamination events (1, 1, 1, 2, and 3 contaminant read-pairs each).

Given these small sample sizes, we expanded our assessment of unintentional contamination rates by applying sentinel-barcoded SIMPLseq to 1420 dried blood spot samples obtained from an efficacy trial of the anti-malaria monoclonal antibody L9LS that was conducted in Mali (16). For the 712 samples (676 pediatric, 36 adult) with parasitemias quantifiable by qRT-PCR (≥ 0.1 p/μl) (Fig. 6), sequencing sensitivity relative to parasitemia was similar to the results obtained from mock dilution curves of cultured parasites. Within the parasitemia interval 0.1 to 0.5 p/μl, 37.4% (61/163) of samples were successfully genotyped at 1 or more loci (median 2 for successful samples). Within the parasitemia interval 0.5 to 1 p/μl, 69.5% (41/59) of samples were successfully genotyped at 1 or more loci (median 3 for successful samples). Genotyping success rose to 89.4% (126/141) for the interval 1 to 10 p/μl (median 5 loci for successful samples) and 98.6% (344/349) for >10 p/μl (median 6 loci for successful samples). These success rates clearly improved on previous attempts (16) to genotype part of the L9LS sample set via 4CAST assay (Supplementary Figure 7).

**Figure 6.**
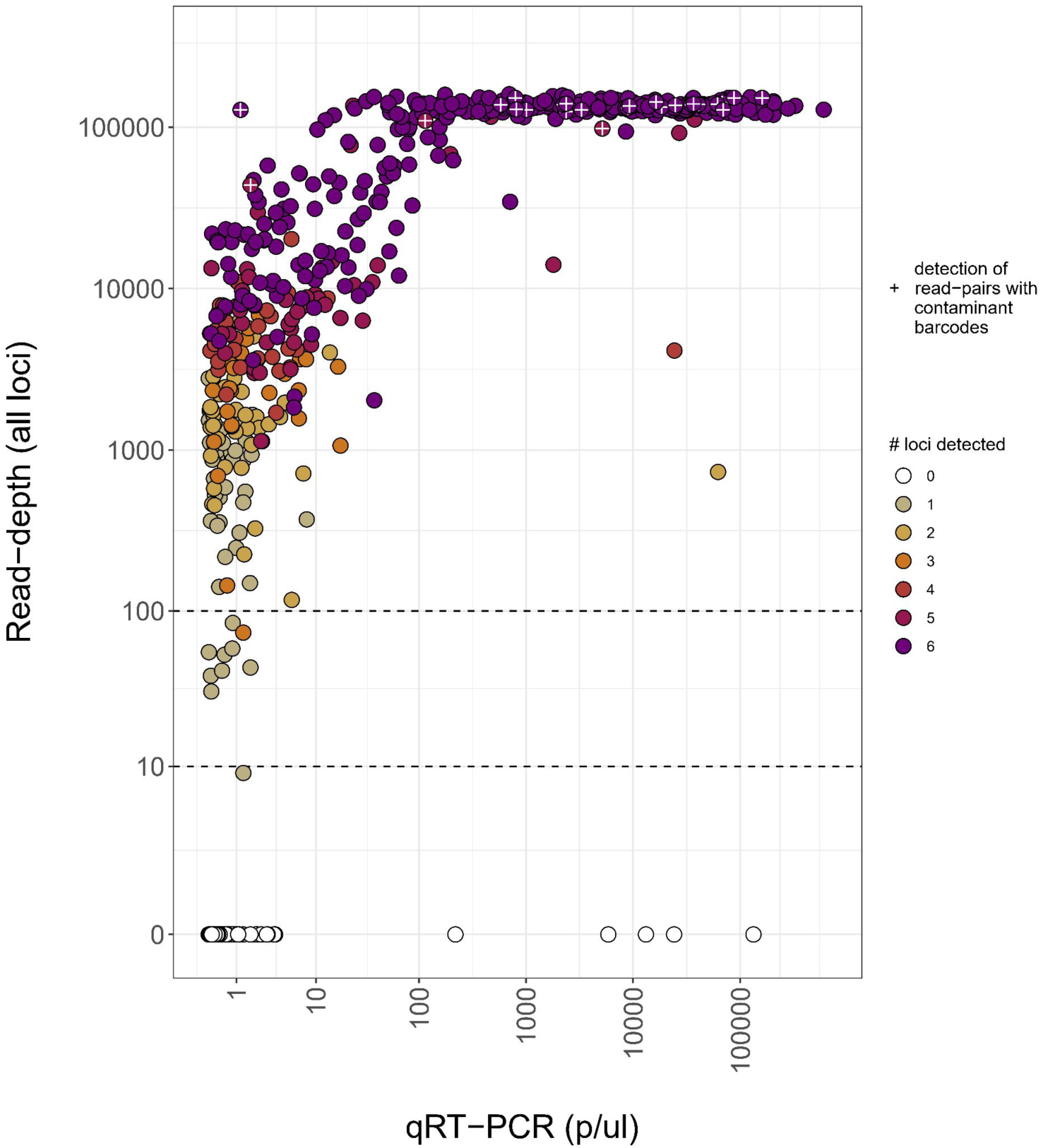
SIMPLseq with sentinel barcoding applied to malaria-infected blood spots collected in Mali (L9LS clinical trial). The plot shows 712 samples (676 pediatric, 36 adult) for which 18S qRT-PCR quantification was performed (see x-axis). Total sample read-depth is shown on the y-axis and point fill color indicates the number of loci amplified. The analysis does not set a minimum depth filter per locus. Plus (‘+’) symbols indicate the detection of read-pairs with contaminant barcodes in 25 samples.

Contaminated barcode signal was detected for 39 of 1420 (2.7%) samples and these occurred on each of the 16 plates used during PCR1 (1, 1, 2, 2, 2, 2, 2, 2, 2, 2, 2, 3, 3, 4, 4, and 5 sample wells per plate). Contaminant barcode signal was very low, ranging from 1 to 3 read-pairs per sample, possibly reflecting very low-level, trace contaminations (e.g., aerosolic primer transfers) as opposed to droplet transfers of gDNA or of amplicons as implemented in our mock contamination experiment. A no-template control additionally showed 9 contaminant read-pairs. This control belonged to PCR1 plate #16, which also featured the highest total number (5) of affected sample wells. Overall, contamination presence (yes/no) was more common in higher-parasitemia samples (odds ratio = 1.82 (95% CI: 1.40 - 2.36, p = 5.95 e-06)) despite their low frequency on the run (e.g., of all clinical trial samples with quantifiable parasitemia, only 34.8% (248/712) exceeded 100 p/μl and 49.0% (349/712) exceeded 10 p/μl) (Fig. 6).

## Discussion

This study introduces a new, 6-locus AmpSeq miniplex called SIMPLseq and explores its potential to detect contamination events via inline-barcoding at PCR1, either across all loci or at a single ‘sentinel’ locus. SIMPLseq maintains the high level of haplotypic diversity established previously via the 4CAST design, outperforming smaller panels (and an AMA1 monoplex) in distinguishing pairs of genetically distinct strains sampled from Pf7k WGS data. In the laboratory, SIMPLseq sensitivity exceeded that of the smaller 4CAST multiplex due to enhanced reaction specificity, yielding interpretable data for sample parasitemias at and below 1 p/μl. Successful genotyping at these low parasitemias is critical for analyzing sample sets in which the majority of infections contain less than 10 p/μl, as was the case for this study’s analysis of the L9LS clinical trial that involved school-aged children in Mali who often have low-level, asymptomatic infections (16).

Importantly, the addition of inline barcodes to the PCR1 primers of a single sentinel locus (KELT) did not compromise sequencing coverage and identified 100% of wells deliberately subjected to contamination scenarios involving PCR1 products. Our results also indicated potential for bioinformatic removal of reads representing PCR1 product contamination at the KELT locus. The enhanced limits of detection and the reliable PCR1 product contamination detection rates we report here help further position AmpSeq for use in important precision-genotyping contexts. Relevant use cases for SIMPLseq include COI estimation as a proxy for transmission intensity, detection of clonal transmission in outbreak settings, and molecular infection endpoint analysis for malaria intervention clinical trials. SIMPLseq uses exclusively open-source reagents and does not require sWGA pre-amplification, making it simpler, less expensive, and yet significantly more sensitive than most other *P. falciparum* targeted sequencing protocols.

The method nevertheless warrants several caveats. Notably, inline barcoding is not designed to detect contaminations occurring prior to the PCR1 cycling step. Synthetic DNA spike-in (SDSI) methods, in which unique template molecules (as opposed to unique primers) are used to mark each sample, might be better applicable at pre-cycling steps, but such early tracking potential has yet to be shown (26). A second caveat we observed with inline barcoding was reduced reaction yield when simultaneously applying inline barcodes to all six primer pairs of the SIMPLseq panel instead of to just one sentinel locus. Ideally, all loci could be barcoded without yield reductions such that cases of contamination could be fully scrubbed (all contaminant reads filtered out) using bioinformatics as opposed to re-running the sample through PCR. Regarding limitations to study design, we should also highlight that our inline barcode tests did not explicitly investigate potential relationships between contamination trackability and template concentration differences between donor and recipient wells. We did however observe a positive post-hoc association between parasitemia (as estimated by qRT-PCR) and contamination signal presence when applying inline-barcoded SIMPLseq to a large clinical sample set (L9LS clinical trial). A 10-fold increase in parasitemia increased the odds of signal presence by 1.81x. One hypothesis is that instances of primer-only contamination (i.e., inconsequential contamination with respect to insert genotyping analysis), are more likely to be detected when recipient wells contain many binding opportunities due to abundant template. The above topics warrant further investigation.

## Conclusions

This study provides four useful lessons broadly relevant to AmpSeq users in malaria research and other fields. First, our work underscores the importance of deliberate primer development towards low off-target binding affinity in the context of multiplex PCR. Unoptimized primer sets that perform adequately at high target DNA concentrations (e.g., 4CAST) may generate substantial nonspecific reaction products once applied to lower-parasitemia sample sets. Depending on size and molarity, non-specific products often remain resistant to bead-based cleanups, reducing sequencing quality or contributing to run failure on patterned Illumina flow cells.

Second, assay sensitivity is generally improved by reducing panel size, and a modest set of diverse targets can outperform a single hyperdiverse locus such as AMA1 in resolving genetically distinct strains, particularly in lower-transmission contexts. Relative to single-locus AmpSeq, multiplexing involves virtually no additional laboratory cost, but it must be verified that incrementing panel size does not compromise sensitivity requirements. Achieving adequate assay sensitivity also requires attention to PCR1 input volume when sample parasitemias are expected to be low. For example, considering a 1 p/μl DBS sample and gDNA extraction efficiency of ca. 20% [refs], a typical 3 μl gDNA input to PCR1 has only ca. 45% chance of containing at least one parasite (Poisson process). This chance increases to ca. 91% if gDNA input is increased to 12 μl.

Third, deliberately accounting for contamination risk is important when applying AmpSeq workflows for precision public health, especially when workflows are designed to maximize sensitivity and throughput. Inline barcodes provide an efficient means of detecting PCR product contamination with minimal added laboratory effort and can be coupled to streamlined tracking pipelines. Conventional contamination control measures nevertheless remain very important.

Finally, AmpSeq should not be applied to samples of unknown parasite positivity status because negative samples may be more susceptible to undetectable contamination events (e.g., from before PCR1 cycling). The broad inclusion of negative and low-parasitemia samples into PCR2 product pools can also bring excessive primer polymers into bead cleanup steps, challenging effective size selection and sequencing success.

These lessons are applicable to all multiplexed AmpSeq panel designs, and will help enhance AmpSeq sensitivity and rigor across the field if incorporated into future large multiplex or miniplex panel designs..

## Data Availability

Sequencing data produced by the study will be available online at NCBI SRA BioProject PRJNA1356533 upon publication.

## Abbreviations

DNA: desoxyribonucleic acid
gDNA: genomic DNA
AmpSeq: amplicon sequencing
PCR: polymerase chain reaction
PCR1: first-round PCR
PCR2: second-round PCR
qRT-PCR: quantitative reverse transcription PCR
SNP: single-nucleotide
INDEL: insertion-deletion
VQSR: variant quality score recalibration
SDSI: synthetic DNA spike-ins
bp: base pairs
kbp: kilobase pairs
p/μl: parasites per microliter
μM: micromolar
v/v: volume/volume
s: seconds
min: minutes
HRP2/3: histidine-rich proteins 2 (PF3D7_0831800) and 3 (PF3D7_1372200)
CSP: circumsporozoite protein (PF3D7_0304600)
AMA1: apical membrane antigen 1 (PF3D7_1133400)
SERA2: serine repeat antigen 2 (PF3D7_0207900)
TRAP: thrombospondin-related anonymous protein (PF3D7_1335900)
WDCP: Gene shorthand as a WD-repeat containing protein (PF3D7_1410300)
KELT: Gene shorthand based on a C-terminal sequence motif (PF3D7_1475900)
SERA8: serine repeat antigen 8 (PF3D7_0207300)
SURFIN 4.2: surface-associated interspersed protein 4.2 (PF3D7_0424400)

## Acknowledgements

This work was supported by an award to D. E. N from the Gates Foundation (INV-052365). The clinical trial in Mali was funded by the National Institute of Allergy and Infectious Diseases. We thank the participants in the clinical trial, the technicians involved in sample processing, and our project managers for study logistics. Thank you especially to Annie Laws, Anne Preston, Jeff Skinner, Shanping Li, Ruchit Panchal, Cheyenne Knox, Selina Bopp, Jonathan Livny, and Emily LaVerriere for help on the project. The contributions of the NIH authors were made as part of their official duties as NIH federal employees, are in compliance with agency policy requirements, and are considered Works of the United States Government. However, the findings and conclusions presented in this paper are those of the authors and do not necessarily reflect the views of the NIH or the U.S. Department of Health and Human Services.

