## Supplementary material for "SIMPLseq: a high-sensitivity *Plasmodium falciparum* genotyping and PCR contamination tracking tool": Supp. Fig. 1

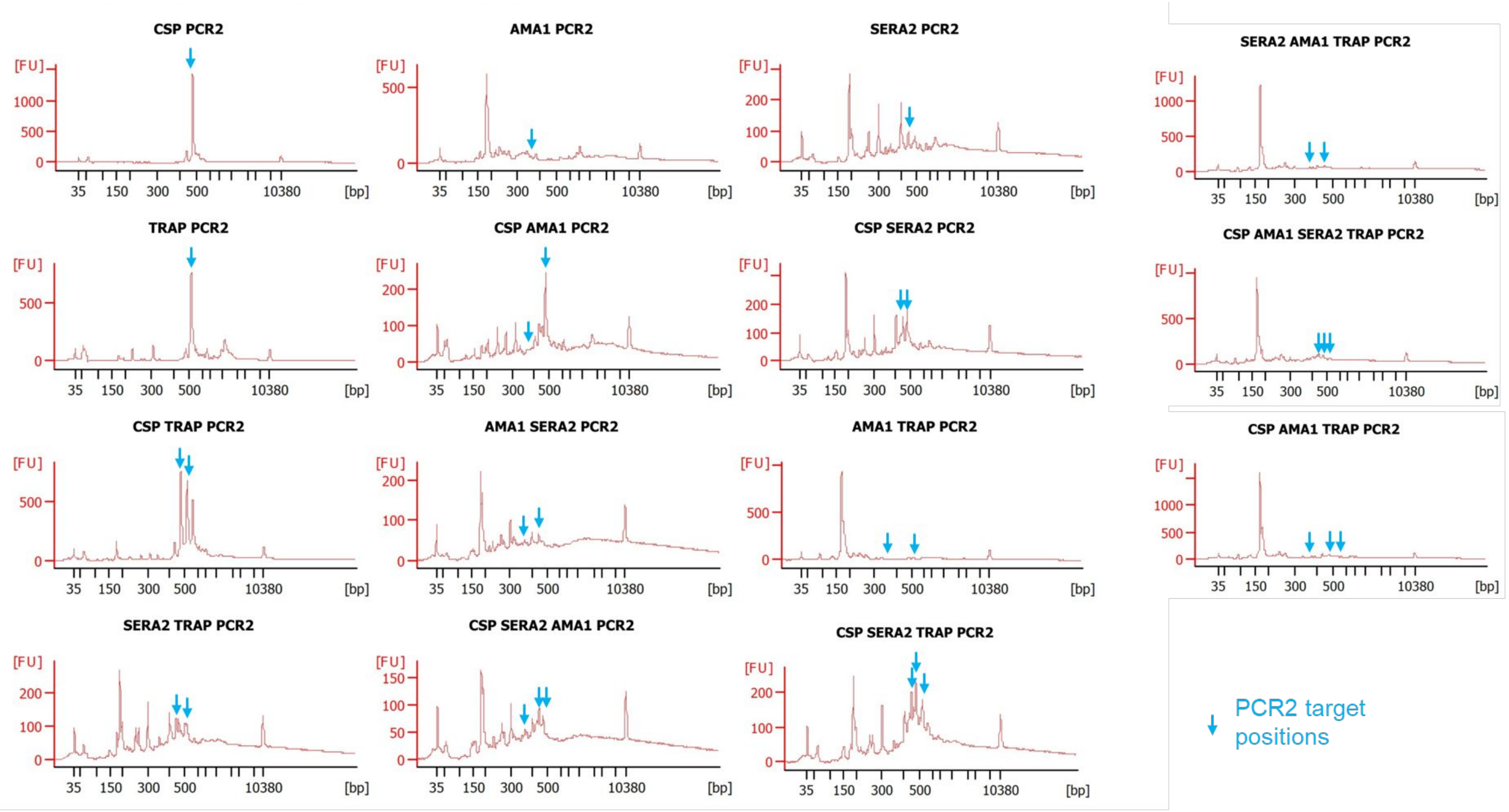


**Supplementary Figure 1. PCR2 product traces for 4CAST and 4CAST subsets prior to bead-cleanup.**
Each trace represents a pool of 5 replicates using 1000 parasites/ul in whole human blood. Note large 190 bp peak and other abundant off-target amplification for reactions involving SERA2 or AMA1 targets. CSP and/or TRAP show higher reaction specificity.
