## Supplementary material for "SIMPLseq: a high-sensitivity *Plasmodium falciparum* genotyping and PCR contamination tracking tool": Supp. Fig. 2

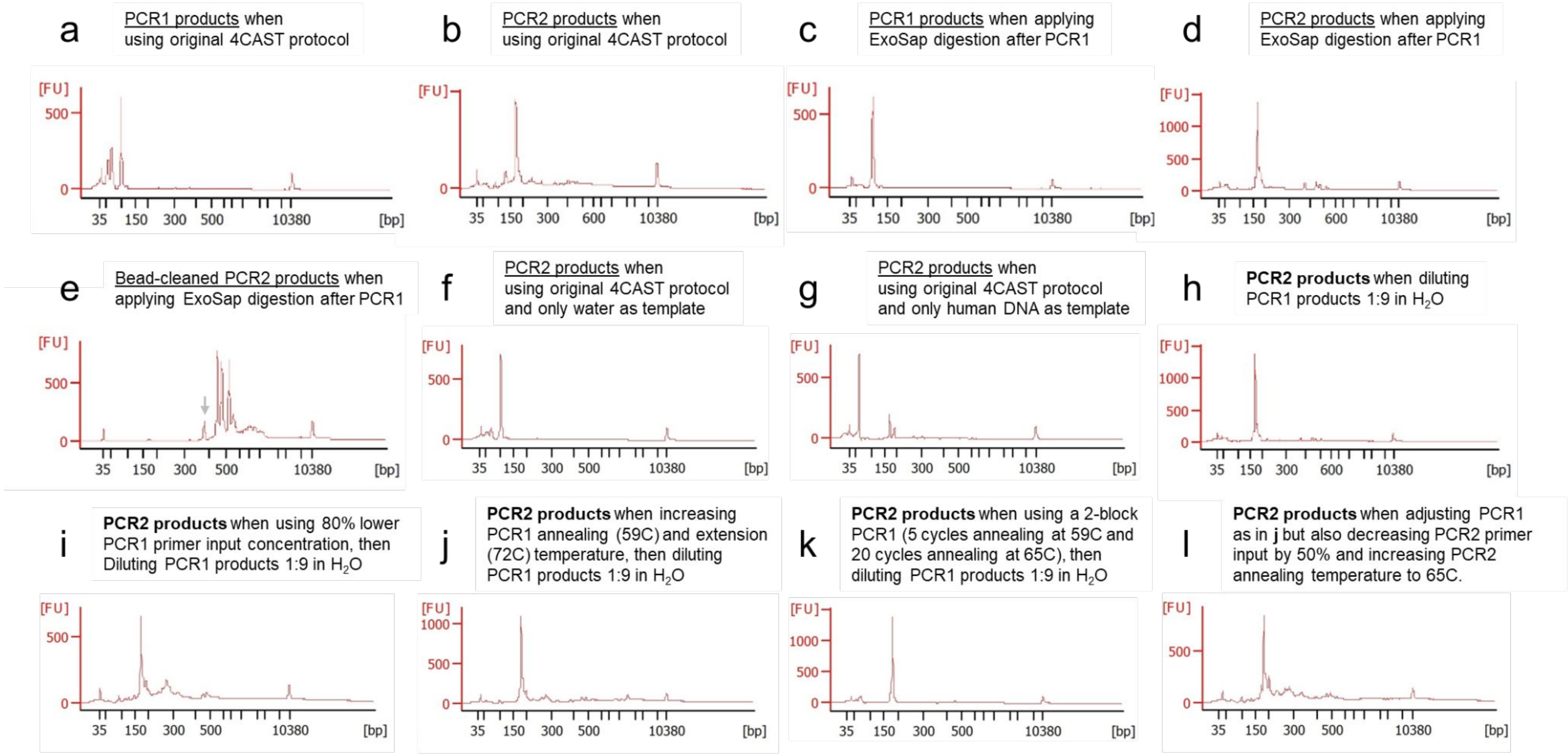


**Supplementary Figure 2. PCR1 and PCR2 product traces (BioAnalyzer) for original and modified 4CAST protocols.** All positive traces represent the use of 16 parasites/ul in whole human blood (5 replicates, pooled) as initial PCR1 input. **a-b**) PCR1 and PCR2 products (prior to bead clean-up) via original 4CAST protocol (LaVerriere et al. 2022). **c-e**) PCR1 and PCR2 products (before and after bead clean-up) when a digestion step (ExoSap) is applied after PCR1. This digestion removes smallest (<75 bp) fragments from PCR1 products and may slightly improve PCR2 target intensities, but off-target products still form abundantly near 190 bp. Off-target clean-up is difficult to achieve without eroding 4CAST targets, especially AMA1 (arrow). **f-g)** Off-target PCR2 products near 190 bp are not as abundant when only water or human DNA is used as PCR1 input. Parasite DNA may therefore be required to generate this artefact. **h-l)** PCR2 products (prior to bead clean-up) for a subset of other attempts (see labels) to improve the original 4CAST protocol. These modifications generally showed inferior clean-up results to those of the protocol involving ExoSap (**e**).
