## Supplementary material for "SIMPLseq: a high-sensitivity *Plasmodium falciparum* genotyping and PCR contamination tracking tool": Supp. Fig. 3

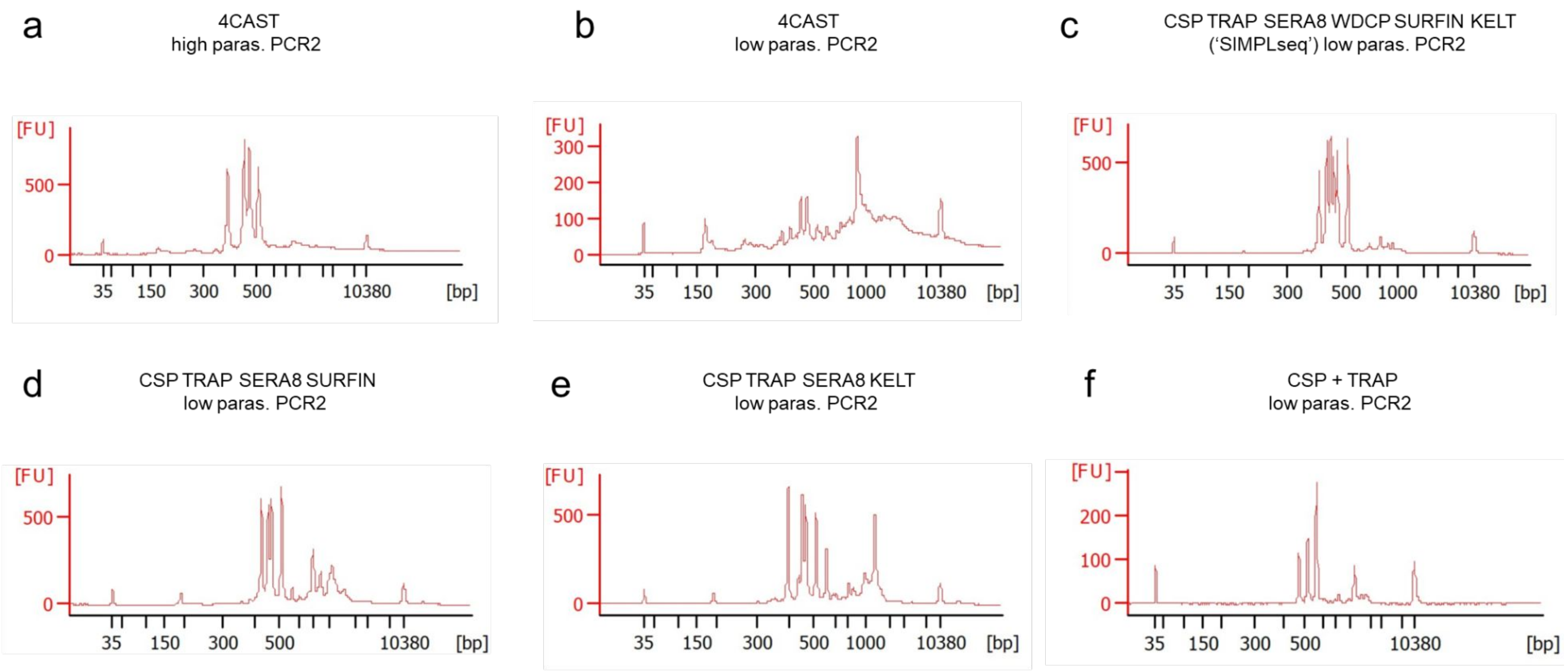


**Supplementary Figure 3. PCR2 product traces for 4CAST and alternative miniplexes after left-sided bead clean-up.**
**a)** 4CAST using 5000 parasites/ul in whole human blood (5 replicates, pooled).
**b-f)** 4CAST and alternative miniplexes using 0.125 – 8 parasites/μl in whole human blood (5 replicates per parasitemia level, pooled).
