## Supplementary material for "SIMPLseq: a high-sensitivity *Plasmodium falciparum* genotyping and PCR contamination tracking tool": Supp. Fig. 4

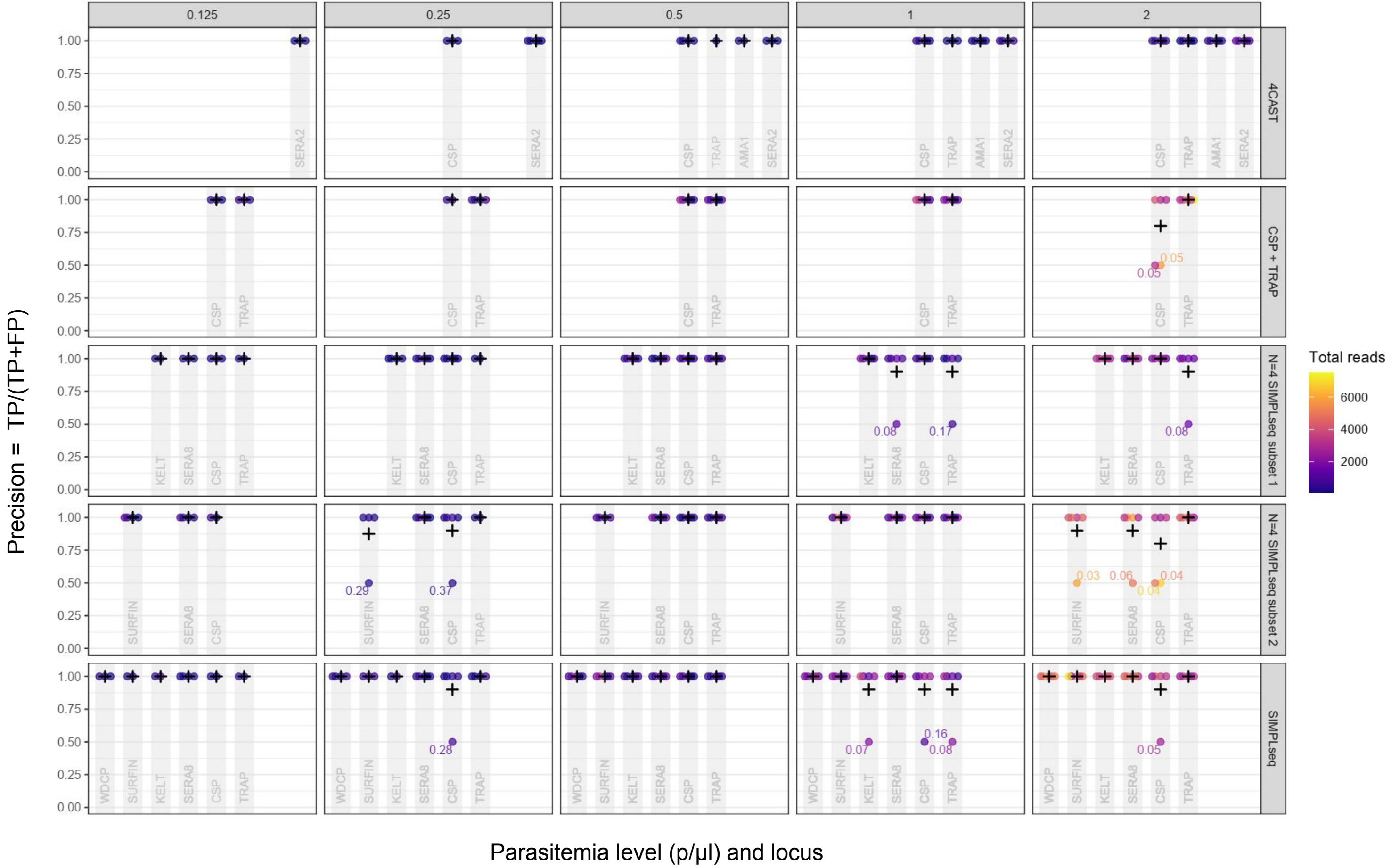


**Supplementary Figure 4. Genotyping precision upon deactivation of the singleton filter.**
Each point represents one of 5 replicates. Y-axis values represent precision (the proportion of detected haplotypes which contained the correct (Dd2-matching) sequence). For example, if a replicate occurs at y-axis value = 0.50, this means there was one additional, incorrect minor haplotype found in that replicate, i.e., 1 of 2 haplotype calls was incorrect; precision was therefore 50% for the replicate.
