## Supplementary material for "SIMPLseq: a high-sensitivity *Plasmodium falciparum* genotyping and PCR contamination tracking tool": Supp. Fig. 5

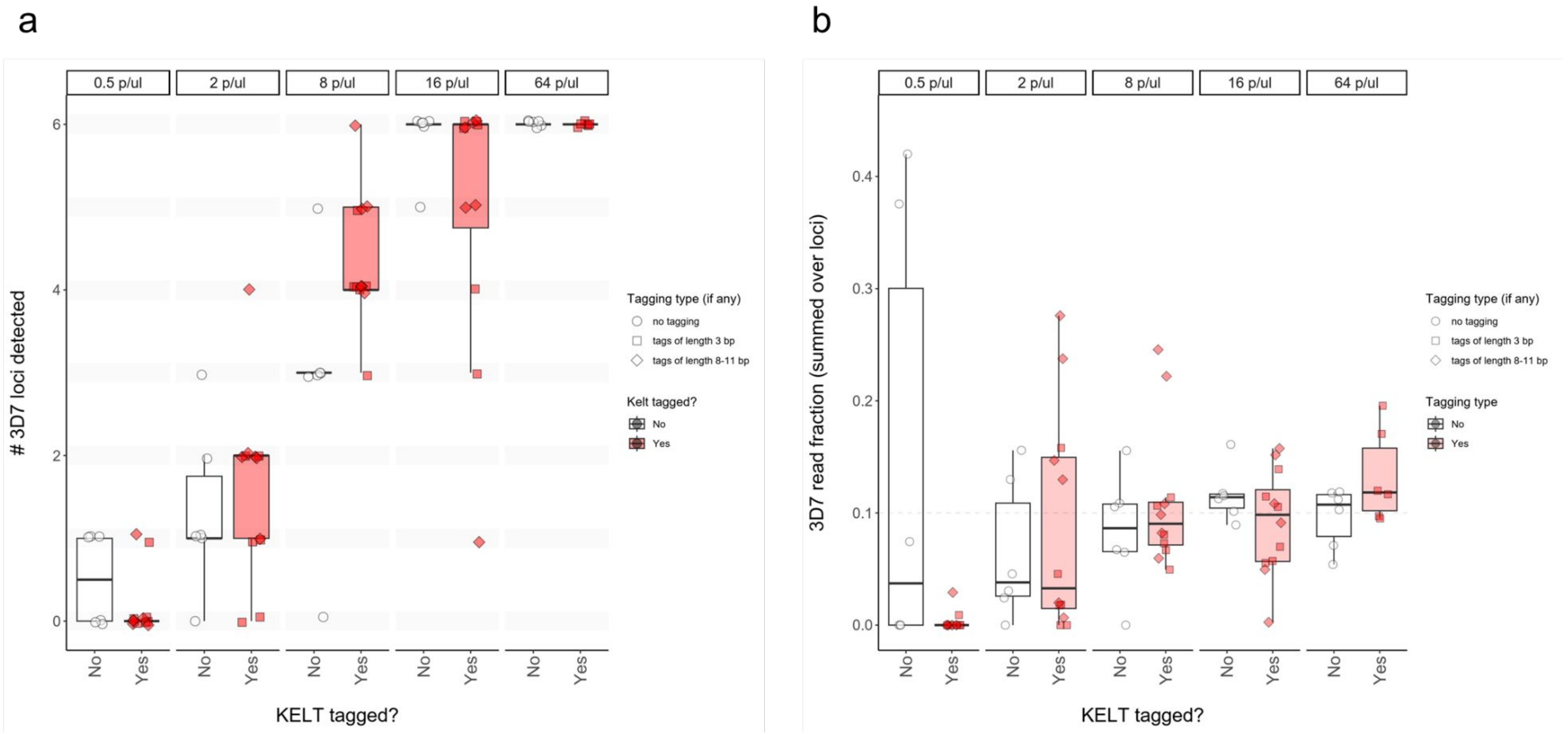


**Supplementary Figure 5. Minor strain detection (untagged and KELT-tagged SIMPLseq applied to mock mixtures of 3D7 and Dd2 parasites).**

Boxplots represent median and interquartile ranges for **a)** the number of 3D7-matching loci and **b)** the 3D7-matching read fraction (i.e., the number of read-pairs matching 3D7 divided by the number of read-pairs matching 3D7 or Dd2, across all loci) when applying SIMPLseq to mock mixtures containing 3D7 (10%) and Dd2 parasites (90%). The x-axis indicates whether the SIMPLseq reaction was untagged or included inline barcoded primer pairs for the KELT locus. For KELT-tagged reactions (see also red fill color), barcodes were either short (3 bp, square symbols) or longer in length (8-11 bp, diamonds). Reactions used parasitemias between 0.5 and 64 p/µl (see facetting left to right). A minimum locus depth filter of 10 was used in the analysis.
