## Supplementary material for "SIMPLseq: a high-sensitivity *Plasmodium falciparum* genotyping and PCR contamination tracking tool": Supp. Fig. 6

| | 1 | 2 | 3 | 4 | 5 | 6 | 7 | 8 | 9 | 10 | 11 | 12 |
|---|---|---|---|---|---|---|---|---|---|---|---|---|
| A | | | | | | | | | | | | |
| B | *GTA/CAT* | *GTA/CAT* | *GTA/CAT* | *GTA/CAT* | *GTA/CAT* | *GTA/CAT* | *GTA/CAT* | *GTA/CAT* | *GTA/CAT* | *GTA/CAT* | *GTA/CAT* | *GTA/CAT* |
| C | ATG/CAT | ATG/CAT | ATG/CAT | ATG/TCA | ATG/TCA | ATG/TCA | ATG/CAT | ATG/CAT | ATG/CAT | ATG/TCA | ATG/TCA | ATG/TCA |
| D | *GTA/CAT* | *GTA/CAT* | *GTA/CAT* | *GTA/CAT* | *GTA/CAT* | *GTA/CAT* | *GTA/CAT* | *GTA/CAT* | *GTA/CAT* | *GTA/CAT* | *GTA/CAT* | *GTA/CAT* |
| E | ATG/CAT | ATG/CAT | ATG/CAT | ATG/TCA | ATG/TCA | ATG/TCA | ATG/CAT | ATG/CAT | ATG/CAT | ATG/TCA | ATG/TCA | ATG/TCA |
| F | *GTA/CAT* | *GTA/CAT* | *GTA/CAT* | *GTA/CAT* | *GTA/CAT* | *GTA/CAT* | *GTA/CAT* | *GTA/CAT* | *GTA/CAT* | *GTA/CAT* | *GTA/CAT* | *GTA/CAT* |
| G | ATG/CAT | ATG/CAT | ATG/CAT | ATG/TCA | ATG/TCA | ATG/TCA | ATG/CAT | ATG/CAT | ATG/CAT | ATG/TCA | ATG/TCA | ATG/TCA |
| H | *ATG/CAT* | *ATG/CAT* | *ATG/CAT* | *ATG/TCA* | *ATG/TCA* | *ATG/TCA* | | | | | | |

| PCR1 input: |
|---|
| 64 p/ul Dd2 |
| 64 p/ul 3D7 |
| H2O |
| no reaction |

**Supplementary Figure 6. Plate layout for the deliberate contamination experiment using an inline-barcoded sentinel locus.**

This plate layout was used to enact 3 different deliberate contamination events:

Event 1: 0.3 µl pipetted from row B into row C just before sealing for PCR1-thermocycle;

Event 2: 0.3 µl pipetted from row D into row E just after PCR1-thermocycle;

Event 3: 0.3 µl pipetted from row F: first, all digestions products correctly pipetted into the PCR2 buffer plate as normal, then an additional transfer of digestion product from row F).

Recipient wells represent either 3D7 (gold fill) or water (white fill) as initial PCR1 input template.

All donor wells represent Dd2 (blue fill) as initial PCR1 input template.

Asterisks indicate wells which were not intended to receive contamination

The inline barcoding used for recipient wells compares to that of donor wells in one of two ways:

Recipient wells in columns 1, 2, 3, 7, 8, and 9 are designated with partially unique barcode pairs with respect to their donor wells, e.g., ATG/CAT vs. GTA/CAT.

Recipient wells in columns 4, 5, 6, 10, 11, and 12 are designated with dually unique barcode pairs with respect to their donor wells, e.g., ATG/TCA vs. GTA/CAT.
