## Supplementary material for "SIMPLseq: a high-sensitivity *Plasmodium falciparum* genotyping and PCR contamination tracking tool": Supp. Fig. 7

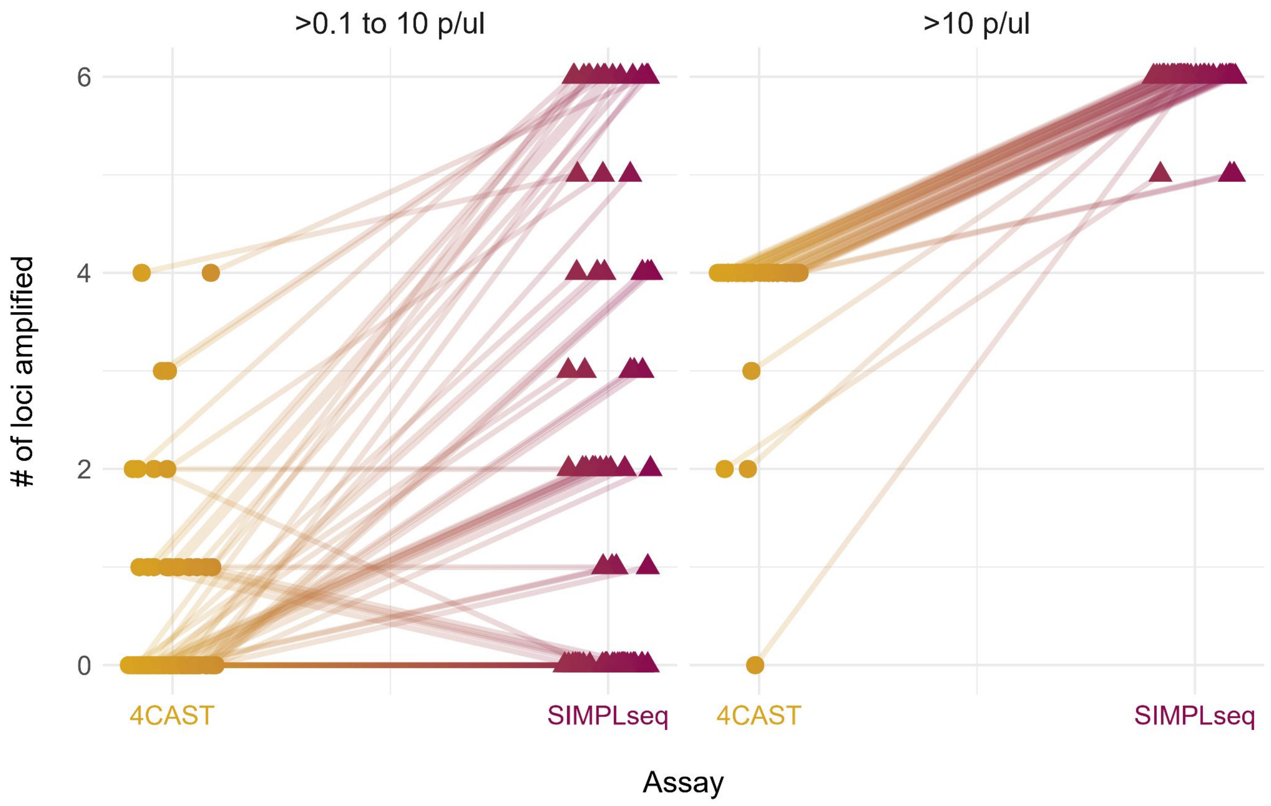


**Supplementary Figure 7. 4CAST versus SIMPLseq (KELT-tagged) application to pediatric cohort samples from Mali.**

4CAST data were obtained for 118 qRT-PCR-positive samples from Kayentao et al. 2024 (NEJM). Each sample was re-assayed using SIMPLseq (including inline barcoded primer pairs for the KELT locus). Lines connect each 4CAST vs.SIMPLseq result (one line per dually-assayed sample). The y-axis indicates the number of detected loci. No minimum locus read-depth threshold is used in this analysis.
